# Sex-specific association of epicardial adipose tissue with coronary artery disease in an Indian cohort: a cross-sectional study

**DOI:** 10.1101/2024.06.12.24308851

**Authors:** Can Xu, Rishabh Khurana, Xuan Gao, Constanze Lehertshuber, Ling Li, Amos Romer, Luigi Filippo Brizzi, Moritz von Scheidt, Anurag Yadav, TBS Buxi, Teresa Trenkwalder, Jason Zhensheng Qu, Dongjin Wang, Zhifen Chen

## Abstract

**Background:** Epicardial adipose tissue (EAT) plays an important role in the pathogenesis of coronary artery disease (CAD). The correlation of EAT volume with CAD or its risk factors, especially the sex-specific correlation, has not been fully characterized. Such a knowledge gap was even larger in the South Asian population given the scarcity of ethnic-specific data. This study intended to evaluate the sex-specific relationship between EAT volume and CAD or its risk factors in an Indian cohort.

**Methods:** The retrospective study included 950 subjects who underwent coronary computed tomography angiography (CCTA) from 2013 to 2016 at Sir Ganga Ram Hospital in India. The EAT volume and CAD status were examined. Values of CAD risk factors were documented for the study subjects, including age, sex, body mass index (BMI), smoking, hypertension, diabetes mellitus, family history of CAD, total cholesterol (TC), low-density lipoprotein cholesterol (LDL-C), and high-density lipoprotein cholesterol (HDL-C) and triglycerides (TG). In a sex-specific fashion, the generalized additive model and multivariable logistic regression analyses were applied to assess the correlation between EAT volume and CAD or its risk factors. The two-piecewise linear regression model was applied to identify the inflection point for the nonlinear correlations.

**Results:** In the 950 subjects, EAT volume was larger in men than in women (67.66 ± 31.83 (n=623) vs 61.93 ± 28.90 (n=327); P = 0.007). After adjusting for confounders, a nonlinear relationship was detected between EAT volume and CAD in the overall subjects and men, but not in women. The inflection point for men was 90ml. The effect sizes and the confidence intervals of EAT volume on CAD were larger when EAT volume was < 90ml. Moreover, we found a linear correlation between EAT volume and BMI in men of the current cohort. In multivariable analysis, either as a continuous or a categorized variable, EAT volume was significantly associated with CAD and BMI by crude, partially adjusted-, and fully adjusted-models in overall subjects and men. Every 1-SD (31.8ml) increase in EAT of men was associated with a higher risk of CAD (odds ratio (OR): 1.76; 95% CI: 1.36 to 2.28; p < 0.00001) by a fully adjusted model. However, EAT volume was not associated with other risk factors. In women of this cohort, EAT volume was not associated with CAD. Interaction analysis indicated BMI influenced the EAT and CAD association specifically in men. EAT volume and CAD showed a stronger association in men with a BMI < 30 kg/m^2^ than ≥ 30 kg/m^2^ (Interaction P=0.0381).

**Conclusion:** EAT volume, an indicator of organ obesity, was positively and independently correlated with CAD in men of the current Indian cohort. In the male subjects, the correlation of EAT volume with CAD was nonlinear, and with BMI was linear. EAT and CAD showed a stronger association in men with EAT volume less than 90 ml or BMI less than 30 kg/m^2^. In women of the current cohort, EAT was not associated with CAD and investigated risk factors, suggesting sex-specific effects of EAT volume on cardiovascular diseases.

## Introduction

Coronary artery disease (CAD) represents the leading cause of death worldwide which is caused by blockages of cardiac blood vessels (coronary arteries) due to the build-up of lipid-rich inflammatory plaques, i.e., atherosclerosis. General obesity, defined by body mass index (BMI ≥30kg/m^2^), is an independent risk factor for CAD. Ectopic fat distribution, visceral fat accumulation, organ obesity, and abdominal or central obesity increase the CAD risk independently of BMI [1]. South Asian countries (India, Pakistan, Nepal, Bhutan, Bangladesh, Sri Lanka, and Maldives) account for approximately ∼25% of the world’s population, yet they contribute >50% of global cardiovascular deaths [2–4]. Of note, India, accounting for ∼76% of the South Asian population [4], is almost as populated as China. According to a recent Nationally Representative Study of India (2023) [5], 13.85% of Indians are obese, doubled since 2008 [6]. Despite the high prevalence, the obesity rate among Indians is approximately one-third less than American and European [7]. However, in India, the prevalence of central obesity (57.71%) is higher than the global average (41.5%) and represents a main driver for obesity-associated CAD in India [5][7–9]. Central obesity, also known as trunk obesity, is known as an excessive accumulation of visceral adipose tissue (VAT) surrounding organs. Epicardial adipose tissue (EAT), a type of VAT, is referred to as the adipose depot beneath the pericardium, and in direct contact with the myocardium and coronary arteries without fibrous fascia separation. Cytokines, adipokines, gaseous molecules, and extracellular vesicles from EAT directly interact with coronary arteries and myocardium via paracrine, endocrine, and vasocrine effects [10]. Proinflammatory EAT was proven to alter vascular tone and promote endothelial dysfunction and progression of cardiovascular diseases (CVDs) [11]. Statin therapy and lifestyle improvement were shown to decrease the accumulation and inflammatory profile of EAT [12–14]. Thus, EAT represents both a predictive maker and a modifiable risk factor for CAD prevention [15].

Recent years have witnessed a growing interest in the relationship between the EAT and CAD risk [16–18]. With the rapid development of imaging technologies, the spatial and temporal resolution of computed tomography scanning has significantly improved in recent years, popularizing its applications in the clinical diagnosis of visceral adipose content and CAD. Coronary computed tomography angiography (CCTA) represents a non-invasive imaging approach to assess and quantify both CAD severity and EAT volume [19]. The correlation between EAT volume and CAD risk has been previously investigated by CCTA and other imaging techniques [20]. However, whether EAT can be considered a predictive marker for CAD occurrence and development remains controversial and the clinical significance of EAT volume on CAD is largely unknown. Of note, ethnic differences in EAT volume have been observed and therefore ethnicity might be considered when EAT volume is used as a predictor for CAD risk [21–23]. While many studies on EAT-CAD correlation were conducted in Western and Eastern Asian cohorts [24,25], such studies in South Asia are scarce. Further, both the associations between EAT volume with CAD risk factors and the sex-specific effect of EAT have not been studied in South Asian cohorts. In the current study, we investigated the sex-specific correlation of EAT volume with not only CAD but also its risk factors in a large Indian clinical imaging cohort.

## Methods

### Study subjects

This analysis utilized data from the prospective cum retrospective cross-sectional observational study from 2013 to 2016 at Sir Ganga Ram Hospital in India, which, by CCTA, examined symptomatic subjects, who were suspicious for CAD but unsuitable for functional tests or with an uninterpretable electrocardiogram. Subjects examined by CCTA were screened for inclusion in the current study. The demographic data, the status of CAD, CAD risk factors (smoking status, family history of CAD, BMI, hypertension, diabetes mellitus, lipid levels, and other CVDs), and the clinical symptoms at the time of the CCTA study were documented. After excluding the individuals with missing values, 950 subjects with parameters of EAT volume, status of CAD and the related risk factors, and lipid levels were retained for our analysis. Hypertension was referred to as treated hypertension, untreated diastolic blood pressure ≥ 90 mm Hg, or untreated systolic blood pressure ≥ 140 mm Hg. Diabetes mellitus was defined as treatment with insulin or oral hypoglycemic medications or fasting glucose ≥ 126 mg/dl. For lipid parameters, the cut-off values for total cholesterol (TC), serum low-density lipoprotein cholesterol (LDL-C), serum high-density lipoprotein cholesterol (HDL-C), and serum triglycerides (TG) were 200 mg/dL, 100 mg/dL, 40 mg/dL, and 150 mg/dl, respectively. Smoking was defined as current or former smokers. The study has been approved by the Institutional Review Board of the Sir Ganga Ram Hospital following the guidelines of the Declaration of Helsinki. All patients provided written informed consent before data collection.

### CCTA-based evaluation of EAT volume and CAD status

At the time of the study, all enrolled subjects were in sinus rhythm, and for individuals with heart rate > 60 beats/min, a β-blocker and/or an anxiolytic was provided. The β-blocker was replaced by a calcium channel blocker in case of intolerance. Sorbitrate Sublingual (2.5 mg) was administered 2 min before the initial scout image. A prospective ECG-triggered scan without a contrast agent was acquired. Retrospective ECG-triggered contrast-enhanced CCTA scan was conducted after intravenous administration of 80-110 ml of 350 mg/ml nonionic water-soluble agent Omnipaque (iohexol, GE Healthcare, Shanghai, China), at a rate of 5.5 ml/s. CCTA scan was acquired between tracheal bifurcation (above the coronary arteries) and the dome of the diaphragm, on a low-dose128-slice MDCT scanner (Ingenuity Philips, Philips Medical System, Netherlands), using voltage 100-120 kVp, current of 400-450mAs (auto modulated), exposure time of 480 to 600 ms, and helical scan with pitch: 0.2. After manually adjusting the field of view, acquired data was reconstructed at various phases of the cardiac cycle, using slice thickness of 0.8 mm and reconstruction increment of 0.4 mm in dedicated soft-tissue kernel settings.

EAT was quantified based on the CCTA end-systolic phase images. A batch film was reconstructed with a 3 mm slice thickness and a 1.5 mm increment. The EAT area was measured between the origin of the left main coronary artery and the cardiac apex. We utilized volume analysis software on the GE Advantage Window workstation to manually delineate the outer contour (the pericardium) and specified the density range of –250 to –30 Hounsfield units (H.U.) to isolate adipose tissue (Fig. 1). CAD absent was defined as no stenosis and CAD as any degree of stenosis in coronary artery segments.

**Figure 1.**
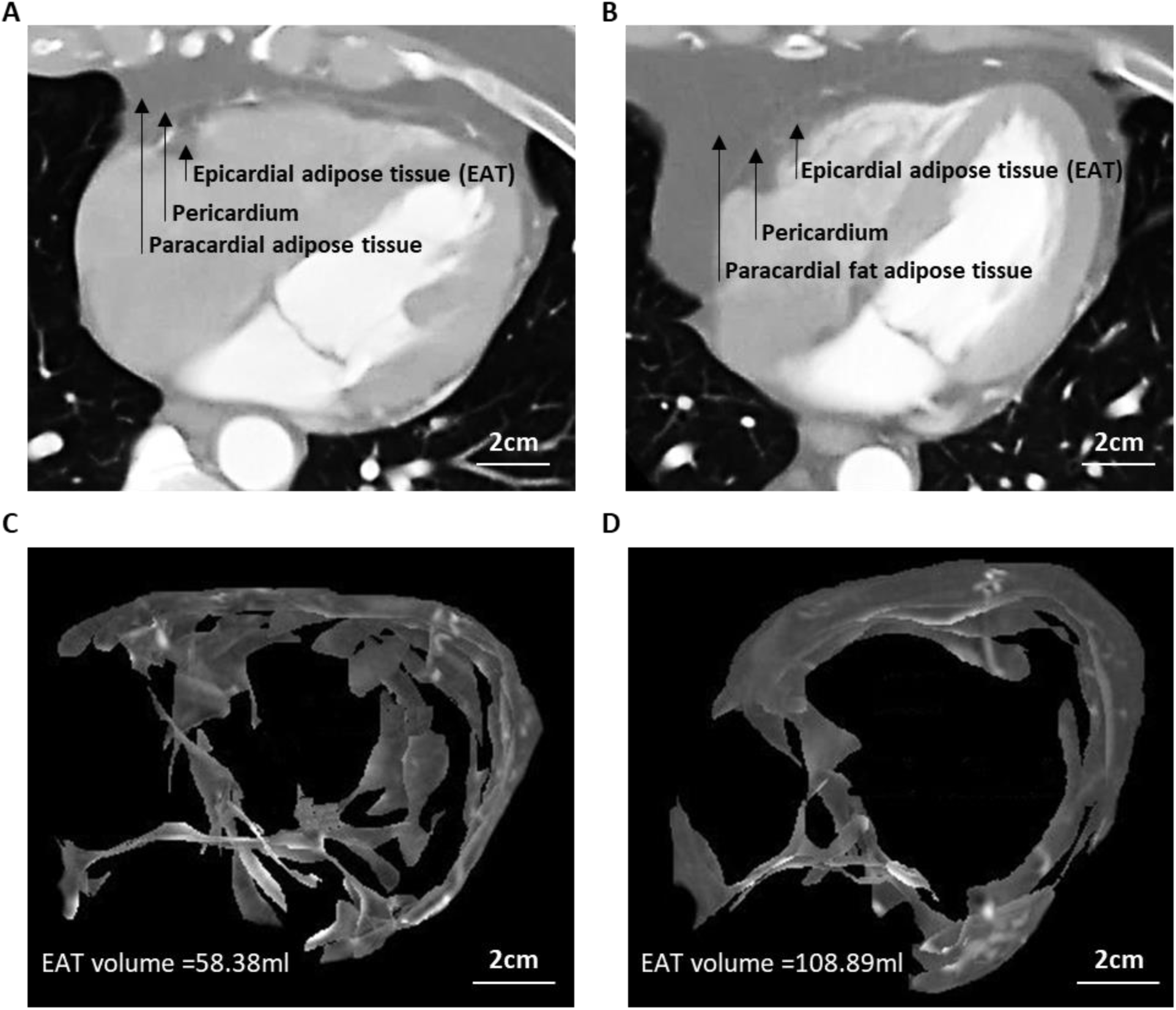
Representative coronary computed tomography angiography (CCAT) images of epicardial adipose tissue (EAT) of a control and a patient with coronary artery disease (CAD). **(A-B)** Thoracic CCAT images of a control **(A)** and a CAD patient **(B)**. EAT, Pericardium, and paracardial adipose tissue are indicated by the arrows. **(C-D)** Isolated EAT fraction of the same control **(C)** and the same CAD patient **(D)**.

### Statistical analysis

Continuous variables were expressed as mean ± standard deviation (normal distribution) or median (interquartile range) (skewed distribution), and categorical variables were presented in frequency or as a percentage. We log-transformed EAT volume and evaluated EAT as continuous (per SD increase) and categorical variables (quantified). The One-Way ANOVA (normal distribution), Kruskal Wallis H (skewed distribution), and chi-square tests (categorical variables) were used to determine any statistical differences between the means and proportions of the groups in a sex-specific manner. We used the generalized additive model to identify the non-linear relationship between EAT volume and CAD risk or other risk factors. If the non-linear correlation was observed, a two-piecewise linear regression model was performed to calculate the threshold effect of the OR on CAD in terms of the smoothing plot. When the ratio between CAD and OR appears obvious in a smoothed curve, the recursive method automatically calculates the inflection point, where the maximum model likelihood will be used. The univariate linear regression model was used to evaluate the associations between EAT volume and CAD incidence. Further, both non-adjusted and multivariable-adjusted models were implemented. According to the recommendation of the STROBE statement, we simultaneously showed the results of crude-, partially- and fully-adjusted analyses. Whether the covariances would be adjusted, was determined by >10% of odds ratio (OR) when added to the model. The subgroup analyses were performed using stratified linear regression models. The modification and interaction of subgroups were inspected by the likelihood ratio test. All the analyses were performed using the statistical software packages R (http://www.R-project.org, The R Foundation, Version 4.2.0) and EmpowerStats (http://www.empowerstats.com, X&Y Solutions, Inc., Boston, MA). P values less than 0.05 (two-sided) were considered statistically significant.

## Results

### Characteristics of the study subjects

A total of 950 subjects were included in the cross-sectional observational study. The baseline statistics of the study subjects are shown in Table 1. In the overall subjects, the mean EAT volume was 65.7 ± 31.0 ml, the mean age was 51.10 ± 11.2 years, and the mean BMI was 26.2 ± 3.5 kg/m^2^. Among the 950 subjects, 623 (65.6%) were men and 118 (12.4%) were smokers. 220 (23.2%) subjects were with a family history of CAD, 400 (42.1%) with hypertension, 203 (21.4%) with diabetes, 218 (22.9%) with high total cholesterol, 211 (22.2%) with high LDL-C, 147(15.5%) with high TG, and 65 (6.8%) with low HDL-C. a . 458 (48.2%) individuals had CAD. Compared to women, men had significantly higher EAT volume (67.7 ± 31.8 vs 61.9 ± 28.9, P < 0.001), larger BMI (27.0 ± 4.2 vs 25.8 ± 3.1, P < 0.001), more smokers (18.6% vs 0.6%, P = 0.007), more individuals with high TG level (17.3% vs 11.9%, P = 0.029) and higher CAD incidence (54.3% vs 36.7%, P < 0.001) (Table 1). The results of the univariate analysis showed that age, sex, smoking, hypertension, diabetes mellitus, family history of CAD, TC, LDL-C, HDL-C, TG, and EAT volume were significantly and positively correlated with CAD in the overall subjects (Table 2).

**Table 1:** Basic characteristics of the study subjects.

| Characteristic | All subjects<br>N = 950 | Men<br>N = 623 (65.6%) | Women<br>N = 327 (34.4%) | P-value |
| --- | --- | --- | --- | --- |
| **EAT volume (mL)** | 950 (65.7 ± 31.0) | 623 (67.7 ± 31.8) | 327 (61.9 ± 28.9) | 0.007 |
| **Age (years)** | 950 (51.1 ± 11.2) | 623 (50.6 ± 11.7) | 327 (52.0 ± 10.2) | 0.071 |
| **Body Mass Index** | 519 (26.2 ± 3.5) | 334 (25.8 ± 3.1) | 185 (27.0 ± 4.2) | <0.001 |
| **Smoking** |  |  |  | <0.001 |
| No | 832 (87.6%) | 507 (81.4%) | 325 (99.4%) |  |
| Yes | 118 (12.4%) | 116 (18.6%) | 2 (0.6%) |  |
| **Hypertension** |  |  |  | 0.432 |
| No | 550 (57.9%) | 355 (57.0%) | 195 (59.6%) |  |
| Yes | 400 (42.1%) | 268 (43.0%) | 132 (40.4%) |  |
| **Diabetes mellitus** |  |  |  | 0.755 |
| No | 747 (78.6%) | 488 (78.3%) | 259 (79.2%) |  |
| Yes | 203 (21.4%) | 135 (21.7%) | 68 (20.8%) |  |
| **Family history CAD** |  |  |  | 0.906 |
| No | 730 (76.8%) | 478 (76.7%) | 252 (77.1%) |  |
| Yes | 220 (23.2%) | 145 (23.3%) | 75 (22.9%) |  |
| **Total cholesterol** |  |  |  | 0.192 |
| < 240 mg/dL | 732 (77.1%) | 472 (75.8%) | 260 (79.5%) |  |
| ≥ 240 mg/dL | 218 (22.9%) | 151 (24.2%) | 67 (20.5%) |  |
| **LDL-C** |  |  |  | 0.551 |
| < 160 mg/dL | 739 (77.8%) | 481 (77.2%) | 258 (78.9%) |  |
| ≥ 160 mg/dL | 211 (22.2%) | 142 (22.8%) | 69 (21.1%) |  |
| **HDL-C** |  |  |  | 0.361 |
| > 40 mg/dL | 885 (93.2%) | 577 (92.6%) | 308 (94.2%) |  |
| ≤ 40 mg/dL | 65 (6.8%) | 46 (7.4%) | 19 (5.8%) |  |
| **Triglycerides** |  |  |  | 0.029 |
| < 200 mg/dl | 803 (84.5%) | 515 (82.7%) | 288 (88.1%) |  |
| ≥ 200 mg/dl | 147 (15.5%) | 108 (17.3%) | 39 (11.9%) |  |
| **CAD** |  |  |  | <0.001 |
| No | 492 (51.8%) | 285 (45.7%) | 207 (63.3%) |  |
| Yes | 458 (48.2%) | 338 (54.3%) | 120 (36.7%) |  |

Values are N (Mean±SD) or N (%). EAT, epicardial adipose tissue; CAD, coronary artery disease; LDL-C, low density lipoprotein cholesterol; HDL-C, high density lipoprotein cholesterol; TG, triglycerides. Hypertension was referred as treated hypertension, or untreated diastolic blood pressure ≥ 90 mm Hg, or untreated systolic blood pressure ≥ 140 mm Hg. Diabetes mellitus was defined as treatment with insulin or oral hypoglycemic medications or fasting glucose ≥ 126 mg/dl.

**Table 2:** Univariate analysis for coronary artery disease.

|  | All subjects (N = 950) |  |  | Men (N = 623 ) |  |  | Women (N = 327) |  |  |
| --- | --- | --- | --- | --- | --- | --- | --- | --- | --- |
| Covariate | Statistics | OR (95%CI) | P-value | Statistics | OR (95%CI) | P-value | Statistics | OR (95%CI) | P-value |
| **EAT volume (mL)** | 65.69 ± 30.96 | 1.96 (1.69, 2.28) | <0.0001 | 67.66 ± 31.83 | 1.95 (1.62, 2.35) | <0.0001 | 61.93 ± 28.90 | 1.92 (1.48, 2.50) | <0.0001 |
| **Age (years)** | 51.10 ± 11.23 | 1.09 (1.08, 1.11) | <0.0001 | 50.62 ± 11.72 | 1.10 (1.08, 1.12) | <0.0001 | 52.00 ± 10.18 | 1.10 (1.07, 1.13) | <0.0001 |
| **Smoking** |  |  | <0.0001 |  |  | 0.001 |  |  | 0.9807 |
| No | 832 (87.58%) | Refrence |  | 507 (81.38%) | Refrence |  | 325 (99.39%) | Refrence |  |
| Yes | 118 (12.42%) | 2.64 (1.75, 3.99) |  | 116 (18.62%) | 2.04 (1.33, 3.14) |  | 2 (0.61%) | NA |  |
| **Hypertension** |  |  | <0.0001 |  |  | <0.0001 |  |  | <0.0001 |
| No | 550 (57.89%) | Refrence |  | 355 (56.98%) | Refrence |  | 195 (59.63%) | Refrence |  |
| Yes | 400 (42.11%) | 2.84 (2.18, 3.71) |  | 268 (43.02%) | 2.62 (1.89, 3.65) |  | 132 (40.37%) | 3.47 (2.17, 5.55) |  |
| **Diabetes mellitus** |  |  | <0.0001 |  |  | <0.0001 |  |  | 0.0021 |
| No | 747 (78.63%) | Refrence |  | 488 (78.33%) | Refrence |  | 259 (79.20%) | Refrence |  |
| Yes | 203 (21.37%) | 2.83 (2.04, 3.93) |  | 135 (21.67%) | 3.30 (2.15, 5.08) |  | 68 (20.80%) | 2.34 (1.36, 4.03) |  |
| **Family history of CAD** |  |  | 0.0008 |  |  | 0.0194 |  |  | 0.0104 |
| No | 730 (76.84%) | Refrence |  | 478 (76.73%) | Refrence |  | 252 (77.06%) | Refrence |  |
| Yes | 220 (23.16%) | 1.69 (1.24, 2.29) |  | 145 (23.27%) | 1.58 (1.08, 2.31) |  | 75 (22.94%) | 1.98 (1.17, 3.35) |  |
| **Total cholesterol** |  |  | 0.0013 |  |  | 0.1847 |  |  | 0.0005 |
| < 240 mg/dL | 732 (77.05%) | Refrence |  | 472 (75.76%) | Refrence |  | 260 (79.51%) | Refrence |  |
| ≥ 240 mg/dL | 218 (22.95%) | 1.65 (1.22, 2.24) |  | 151 (24.24%) | 1.29 (0.89, 1.86) |  | 67 (20.49%) | 2.63 (1.52, 4.55) |  |
| **LDL-C** |  |  | 0.0072 |  |  | 0.1827 |  |  | 0.0071 |
| < 160 mg/dL | 739 (77.79%) | Refrence |  | 481 (77.21%) | Refrence |  | 258 (78.90%) | Refrence |  |
| ≥ 160 mg/dL | 211 (22.21%) | 1.53 (1.12, 2.08) |  | 142 (22.79%) | 1.29 (0.89, 1.89) |  | 69 (21.10%) | 2.10 (1.22, 3.59) |  |
| **HDL-C** |  |  | 0.0003 |  |  | 0.0156 |  |  | 0.0057 |
| > 40 mg/dL | 885 (93.16%) | Refrence |  | 577 (92.62%) | Refrence |  | 308 (94.19%) | Refrence |  |
| ≤ 40 mg/dL | 65 (6.84%) | 2.78 (1.60, 4.82) |  | 46 (7.38%) | 2.26 (1.17, 4.39) |  | 19 (5.81%) | 4.07 (1.50, 11.01) |  |
| **Triglycerides** |  |  | <0.0001 |  |  | 0.0006 |  |  | 0.0469 |
| < 200 mg/dl | 803 (84.53%) | Refrence |  | 515 (82.66%) | Refrence |  | 288 (88.07%) | Refrence |  |
| ≥ 200 mg/dl | 147 (15.47%) | 2.21 (1.54, 3.19) |  | 108 (17.34%) | 2.18 (1.40, 3.40) |  | 39 (11.93%) | 1.98 (1.01, 3.88) |  |

Values are N (Mean±SD) or N (%). EAT, epicardial adipose tissue; CAD, coronary artery disease; LDL-C, low density lipoprotein cholesterol; HDL-C, high density lipoprotein cholesterol; TG, triglycerides; OR, odds ratio; CI, confidence interval.

### Nonlinear correlation of EAT volume with CAD risk in men

Using EAT volume as a continuous variable in the generalized additive model, we visualized the correlation between EAT volume and CAD in the overall subjects, women and men, respectively (Fig. 2). In the overall subjects, we found that the relationship between EAT volume and CAD was non-linear after adjusting for age, sex, smoking, hypertension, diabetes mellitus, family history of CAD, TG, TC, LDL-C and HDL-C (Fig. 2A, Table 3). A similar non-linear correlation in men was observed, but not in women (Fig. 2B, 2C). In women, the correlation of EAT volume with CAD was trend in linear but without significance (Fig. 2C), suggesting the non-linear correlation in overall subjects was driven by men.

**Figure 2.**
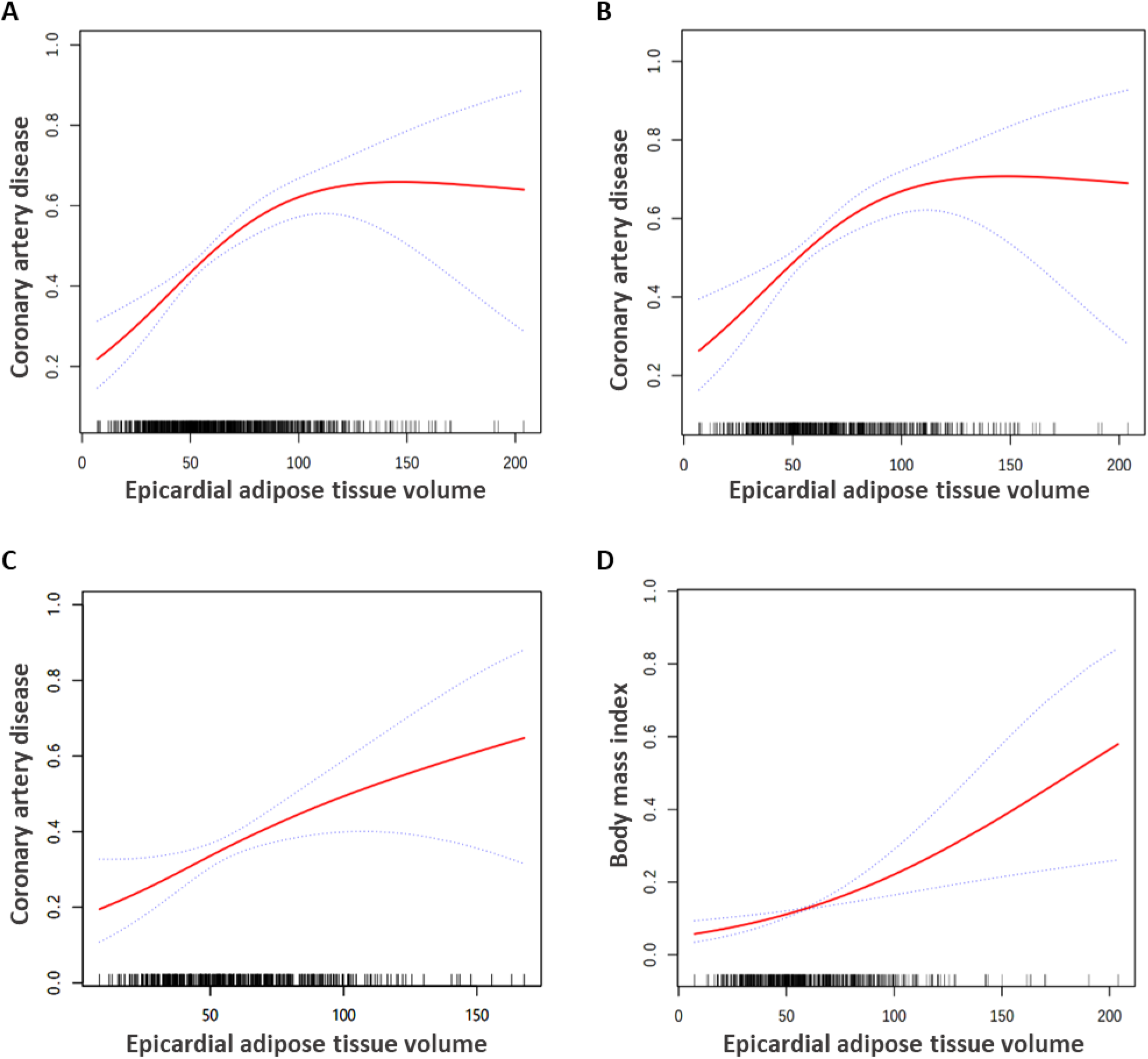
Association between epicardial adipose tissue (EAT) volume and coronary artery disease (CAD) or its risk factors. The generalized additive model (GAM) identified a nonlinear association of EAT volume with CAD in overall subjects **(A)** and in men **(B),** but not in women **(C).** GAM determined a linear correlation between EAT volume and BMI in men **(D).** Among the covariates (age, gender, body mass index, smoking, triglycerides, hypertension, diabetes mellitus, family history CAD, total cholesterol, low­density lipoprotein cholesterol, and high-density lipoprotein cholesterol), except the tested outcome, the rest covariates were adjusted correspondingly. The red solid lines show the estimated risk, and the purple dotted lines represent pointwise 95% Cl.

**Table 3:** Identification of inflecton point by two-piecewise linear regression model.

| The inflection point of EAT volume for CAD in overall subjects |  |  |  |
| --- | --- | --- | --- |
| The inflection point of EAT | Effect size (OR) | 95%CI | P value |
| < 90 ml | 2.32 | 1.88 to 2.87 | < 0.0001 |
| ≥ 90 ml | 1.23 | 0.82 to 1.84 | 0.3099 |

| The inflection point of EAT volume for CAD in men |  |  |  |
| --- | --- | --- | --- |
| The inflection point of EAT | Effect size (OR) | 95%CI | P value |
| < 90 ml | 2.42 | 1.85 to 3.17 | < 0.0001 |
| ≥ 90 ml | 1.12 | 0.70 to 1.79 | 0.6293 |

EAT, epicardial adipose tissue; CAD, coronary artery disease; LDL-C, low density lipoprotein cholesterol; HDL-C, high density lipoprotein cholesterol; TG, triglycerides; OR, odds ratio; CI, confidence interval.

By applying the two-piecewise linear regression model, we found that the EAT volume inflection points for CAD were 90ml in both the overall subjects and men (Fig.2A and 2B, Table 3). In men, on the right of the CAD-EAT volume inflection point (EAT volume ≥ 90 ml), the correlation between EAT volume and CAD was not significant, and the effect size (OR), 95% CI and P value were 1.12, 0.70-1.79 and 0.6293, respectively (Fig.2B, Table 3). Interestingly, we observed a significant and positive correlation between EAT volume and CAD on the left side of the inflection point (< 90 ml) with more than doubled effect size, OR =2.42, 95% CI =1.85 to 3.17, and P value < 0.0001 (Fig.2B, Table 3). In the overall subjects, the results were similar to that of men (Fig.2A, Table 3).

### Sex-specific association of EAT volume with CAD risk

By the multivariable logistic regression analysis, the associations between EAT volume and CAD risk were further investigated using EAT volume as both a continuous and a categorized variable in a sex-specific manner (Table 4). Three models were used: crude, partially-adjusted (adjusted for age), and fully-adjusted (adjusted for age, sex, smoking, hypertension, diabetes mellitus, family history of CAD, TG, TC, LDL-C, and HDL-C). In the sex-specific analysis, the sex was not adjusted in the third model. As a continuous variable, EAT volume was significantly associated with CAD in the overall subjects and in men by all three models (Fig. 3), but not in women, suggesting a sex-specific effect. In the partially-adjusted model, every 1-SD increment in EAT volume increased the risk of CAD by 54% (95% CI: 31% to 82%; p < 0.001) in the overall subjects and 62% (95% CI: 27% to 107%; p < 0.001) in men (Table 4). Additional adjustment for variables in fully adjusted models increased the odds ratio (OR) from 1.54 (95% CI: 1.31 to 1.82; P < 0.0001) to 1.56 (1.31 to 1.85, P < 0.0001) in the overall subjects and 1.67 (1.23 to 2.10, P = 0.00012) to 1.76 (1.36 to 2.28, P = 0.00002) in men.

**Figure 3.**
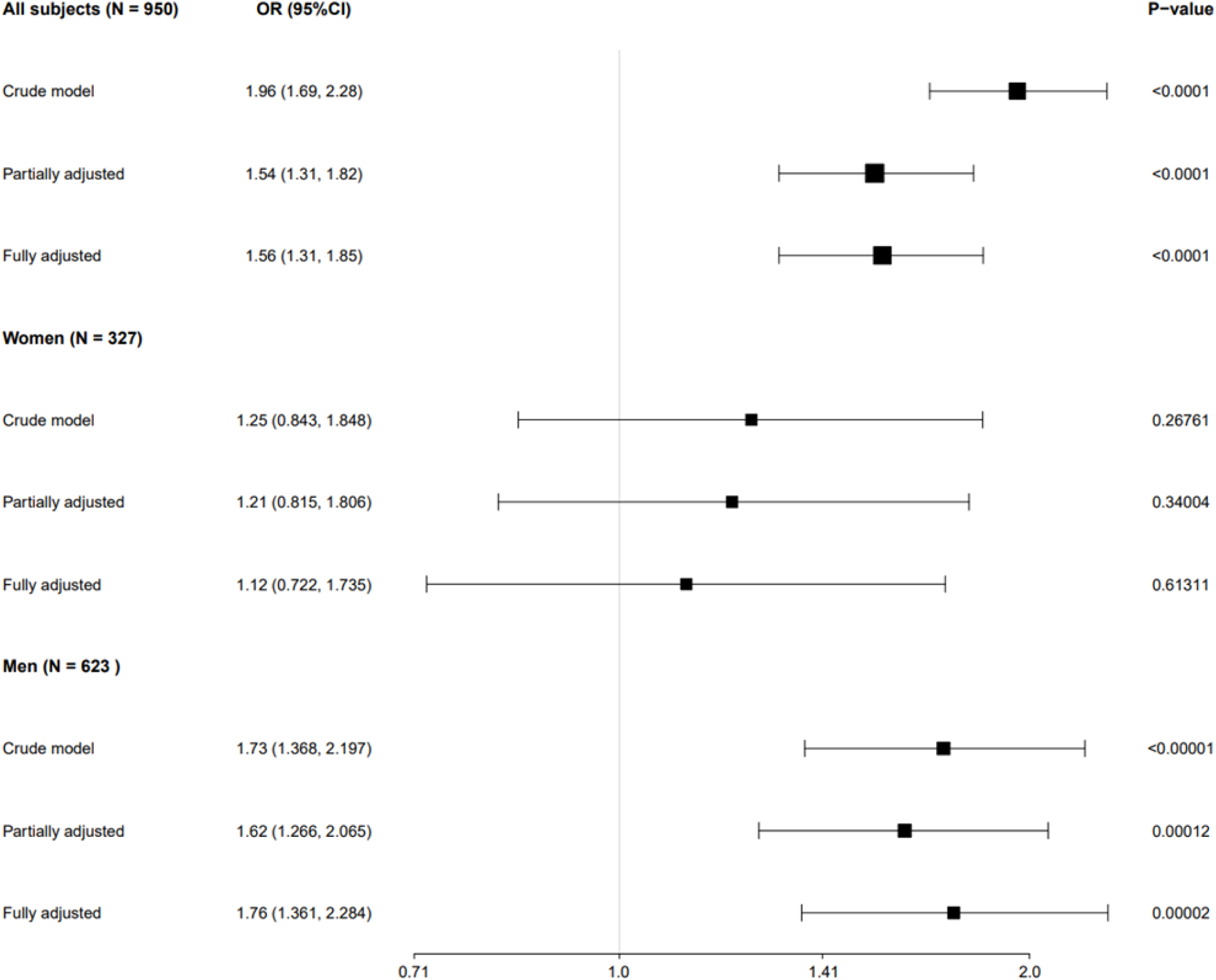
The association EAT volume and CAD risk by multivariable logistic regression analysis. In the partially-adjusted model, age was adjusted. In fully-adjusted model, age, smoking, hypertension, diabetes mellitus, family history of CAD, TG, TC, LDL-C and HDL-C were adjusted. Horizontal error bars denote 95% confidence intervals from the data using EAT as the continuous variable (Table 4).

**Table 4:** Multivariate analyses evaluating the association between EAT volume and the risk of CAD.

| **EAT volume and CAD association in overall subjects** |  |  |  |
| --- | --- | --- | --- |
| **EAT as continuous variable (per 1-SD increment)** |  |  |  |
| Exposure | Crude model | Partially adjusted model | Fully adjusted model |
| EAT volume | 1.96 (1.69, 2.28) <0.0001 | 1.54 (1.31, 1.82) <0.0001 | 1.56 (1.31, 1.85) <0.0001 |
| **EAT as a categorical variable** |  |  |  |
| EAT volume quartile |  |  |  |
| Q1 | 1.00 (referent) | 1.00 (referent) | 1.00 (referent) |
| Q2 | 2.76 (1.87, 4.08) <0.0001 | 1.89 (1.23, 2.91) 0.0039 | 1.74 (1.10, 2.74) 0.0173 |
| Q3 | 3.75 (2.54, 5.54) <0.0001 | 2.33 (1.51, 3.58) 0.0001 | 2.26 (1.44, 3.54) 0.0004 |
| Q4 | 5.67 (3.81, 8.43) <0.0001 | 3.13 (2.02, 4.85) <0.0001 | 3.11 (1.97, 4.93) <0.0001 |
| P for trend | <0.0001 | <0.0001 | <0.0001 |
| **EAT volume and CAD association in men** |  |  |  |
| **EAT as continuous variable (per 1-SD increment)** |  |  |  |
| Exposure | Crude model | Partially adjusted model | Fully adjusted model |
| EAT volume | 1.73 (1.37, 2.20) <0.00001 | 1.62 (1.23, 2.10) 0.00012 | 1.76 (1.36, 2.28) 0.00002 |
| **EAT as a categorical variable** |  |  |  |
| EAT volume quartile |  |  |  |
| Q1 | 1.00 (referent) | 1.00 (referent) | 1.00 (referent) |
| Q2 | 2.39 (1.04, 5.50) 0.04082 | 2.25 (0.96, 5.27) 0.06246 | 2.40 (0.97, 5.92) 0.05695 |
| Q3 | 3.44 (1.53, 7.71) 0.00279 | 3.06 (1.34, 7.00) 0.00793 | 3.67 (1.53, 8.81) 0.00356 |
| Q4 | 6.87 (3.06, 15.42) <0.00001 | 5.91 (2.59, 13.49) 0.00003 | 7.55 (3.14, 18.14) <0.00001 |
| P for trend | <0.00001 | <0.00001 | <0.00001 |
| **EAT volume and CAD association in women** |  |  |  |
| **EAT as continuous variable (per 1-SD increment)** |  |  |  |
| Exposure | Crude model | Partially adjusted model | Fully adjusted model |
| EAT volume | 1.25 (0.84, 1.85) 0.26761 | 1.21 (0.82, 1.81) 0.34004 | 1.12 (0.72, 1.74) 0.61311 |
| **EAT as a categorical variable** |  |  |  |
| EAT volume quartile |  |  |  |
| Q1 | 1.00 (referent) | 1.00 (referent) | 1.00 (referent) |
| Q2 | 1.80 (0.47, 6.87) 0.38955 | 1.52 (0.39, 5.96) 0.55205 | 1.72 (0.34, 8.62) 0.50849 |
| Q3 | 1.65 (0.43, 6.36) 0.46711 | 1.43 (0.36, 5.66) 0.61234 | 1.46 (0.30, 7.16) 0.63935 |
| Q4 | 1.73 (0.47, 6.34) 0.40742 | 1.55 (0.41, 5.79) 0.51748 | 1.41 (0.30, 6.61) 0.65972 |
| P for trend | 0.57901 | 0.63711 | 0.93151 |

In the partially-adjusted model, age (for every 1-year increase) was adjusted. In the fully-adjusted model, age, smoking (no, past, and current), hypertension, diabetes mellitus, family history of CAD, TG, TC, LDL-C and HDL-C were adjusted. 1-SD increment = 31.0 ml, 31.8 ml and 28.9 ml for overall, men and women subjects.

As the categorized variables, the study subjects were divided into four groups according to EAT volume quartile (Q1-Q4), namely Q1 (7.0-43.6ml), Q2 (43.7-59.5ml), Q3 (59.7-82.5ml) and Q4 (82.7-203.9ml) (Supp Table 1). Participants with high EAT volume were older and more males and had higher levels of TC, LDL-C, and TG, and higher prevalence of hypertension, diabetes, and CAD (Supp Table 1). As a categorized variable, an increase in EAT volume was associated with CAD by the crude model in the overall subjects (Table 4). Using Q1 as the reference, EAT volume of Q2-Q3 subjects had an increased effect on CAD with OR=2.76 (1.87 to 4.08; P < 0.0001), 3.75 (2.54 to 5.54; P < 0.0001) and 5.67 (3.81 to 8.43, P < 0.0001) respectively. Further adjustment reduced the odd ratio, but the effect remained significant. When this analysis was independently conducted in men and women (Supp Table 2 and 3), again only in men, the association was significant (Table 4), further confirming the sex-specific association. In men, EAT volume as a categorical variable, comparing with Q1 subjects, Q2-Q3 subjects were respectively associated with 2.39 (1.04 to 5.50; P = 0.04082), 3.44 (1.53 to 7.71; P = 0.00279) and 6.87 (3.06 to 15.42; P < 0.00001) folds of risk increase by the crude model, and even greater risk by the fully-adjusted model, with 2.40 (0.97 to 5.92; P < 0.05695), 3.67 (1.53 to 8.81; P< 0.00356) and 7.55 (3.14 to 18.14; P <0.00001) folds respectively (Table 4).

### Sex-specific effect of BMI on the association of EAT volume and CAD risk

We further tested the associations between EAT volume and CAD risk factors by multivariable logistic regression analysis in a sex-specific manner, including age, sex, smoking, BMI, hypertension, diabetes mellitus, family history of CAD, TG, TC, LDL-C, and HDL-C. In the partially adjusted, age was adjusted, except when age was as the tested variate in the analysis. In the fully adjusted model, except for the tested variate, the rest covariates were adjusted correspondingly. In addition to CAD, EAT volume was significantly associated with BMI, not other risk factors and the significance was exclusive to the overall subjects and men. Similar to the previous correlation between EAT volume and the risk of CAD, EAT volume was significantly associated with BMI by all three models in the overall subjects and men. Every 1-SD increment in EAT volume increased the BMI by 1.84 folds (95% CI: 1.41 to 2.40; p < 0.001) in the overall subjects and 2.12 folds (95% CI: 1.50 to 2.98; p < 0.001) in men by the partially adjusted model (Fig.4, Supp Table 4). Additional adjustment for variables in fully adjusted models decreased the OR from 1.84 (1.41 to 2.40; P < 0.0001) to 1.71 (1.29 to 2.27; P = 0.0002) in the overall subjects and 2.12 (1.50 to 2.98; P < 0.0001) to 1.74 (1.20, 2.52; P = 0.0036) in men (Fig.4, Supp Table 4).

**Figure 4.**
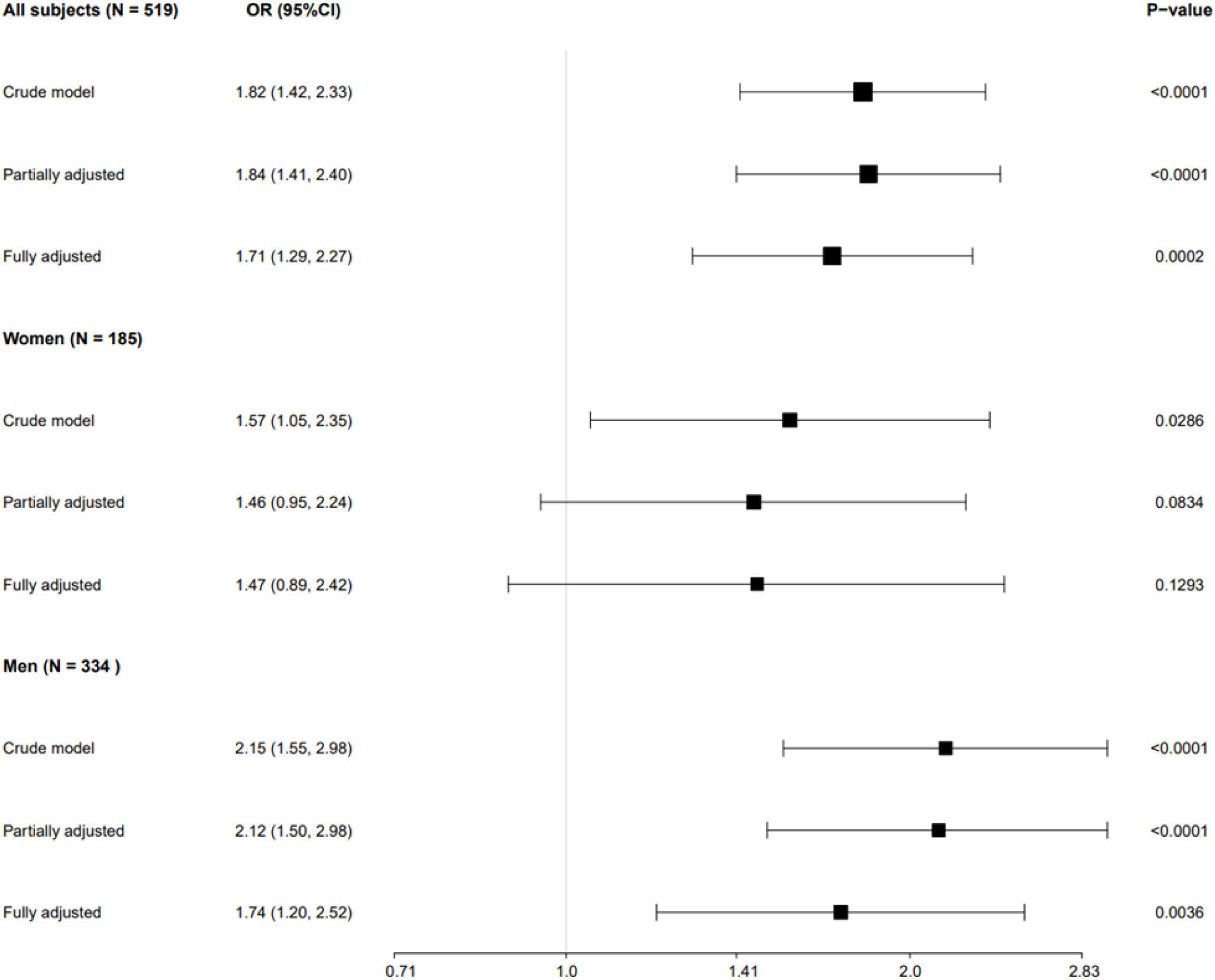
The association of EAT volume and BMI by multivariable logistic regression analysis. In the partially-adjusted model, age was adjusted. In fully-adjusted models, age, smoking, hypertension, diabetes mellitus, family history of CAD, TG, TC, LDL-C and HDL-C were adjusted. Horizontal error bars denote 95% confidence intervals from the data using EAT as the continuous variable (Supplementary Table 4).

When the overall subjects and women were stratified by any of the studied risk factors, the odds ratio between EAT volume and CAD remained similar (Supp Fig.1 and 2). Namely, as shown in Supp Fig.1 and 2, the interaction tests for any of the stratifications were not statistically significant and the interaction P values for age, BMI, sex, smoking, hypertension, diabetes mellitus, family history CAD, TC, LDL-C, HDL-C, TG, respectively were all larger than 0.1 (Supp Fig.1 and 2). Of note, in men, the subgroup analysis of BMI was significant (P=0.0381). EAT volume showed a stronger association with CAD in men with BMI < 30 than those with BMI ≥ 30 (Fig.5), which was not observed in women. This was in line with previous data, namely, EAT showed a strong correlation with CAD in men when the volume was smaller (< 90ml) (Fig. 2, Table 3). In fact, EAT volume had a linear correlation with BMI in men (Fig 2D). These results suggested that BMI influenced the association of EAT volume and CAD specifically in men of the current South Asian cohort.

**Figure 5.**
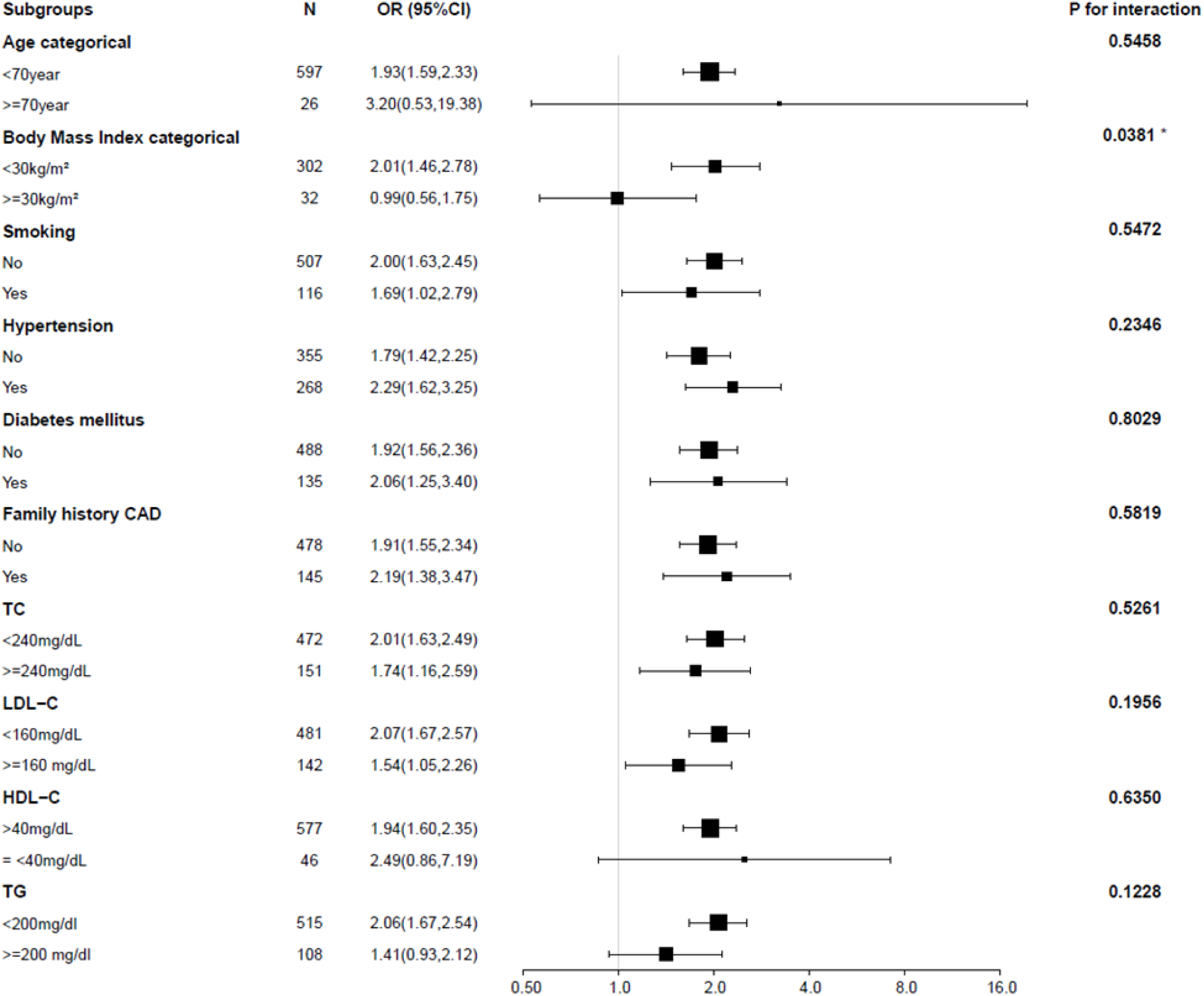
The association of EAT volume and CAD risk according to subgroups in men. The effect of EAT volume on the risk of CAD was similar in various subgroups. All analyses were adjusted for age, smoking, BMI, hypertension, diabetes mellitus, family history of CAD, TG, TC, LDL·Cand HDL·C. Risk estimates were for every 1-SD increase (31.8 ml) in EAT volume. The size of squares is proportional to CAD prevalence in the specified subgroup. Horizontal error bars indicate 95% confidence intervals. N=subjects in each subgroup. OR, odds ratio.

## Discussion

In the present study, we used generalized linear and additive models to elucidate the relationship between EAT volume and CAD or its risk factors in a sex-specific fashion in 950 Indian subjects. Firstly, compared to non-CAD subjects, CAD patients had higher levels of EAT volume, TC, LDL-C, and TG, but a lower level of HDL-C, were composite of more smoking, hypertensive and diabetic subjects, and were more likely to have a family history of CAD (Table 2). CAD prevalence, age, and BMI increased with quantiles of EAT volume (Supp Table 1-3). Secondly, we explored potential curvilinear relationships between EAT volume and CAD or its risk factors in both women and men. By generalized additive model, we found EAT volume was linearly correlated with BMI but nonlinearly correlated with CAD in overall and male subjects, but not in women (Fig.2, Table 3). The inflection points of EAT-CAD correlations in overall and male subjects were 90ml (Table 3). Thirdly, in the crude, partially-, or fully-adjusted models, EAT volume was significantly associated with CAD and BMI in overall and male subjects, but not in women (Fig 3 and 4, Table 4, Supp Table 4). The EAT-CAD associations in overall and woman subjects did not interact with other traditional risk factors of CAD. However, in men, BMI affects the association of EAT volume and CAD risk, especially in men with BMI < 30 kg/m^2^ (Fig.5). These data suggested that in men with BMI < 30 kg/m^2^, visceral obesity (indicated here by large EAT, or ‘heart obesity’) play a stronger role in CAD than general obesity (indicated by large BMI). Conversely, in men with BMI ≥ 30 kg/m^2^, the large BMI might attenuate the effect of EAT on CAD.

### EAT volume correlated with CAD and other cardiometabolic phenotypes

The association between EAT volume and CAD is still debatable [26]. Tanami et al analyzed 320 patients with suspected CAD undergoing 320-detector row CT angiography and found no correlation between EAT volume and the presence or severity of CAD [27]. Likewise, Yin R et al enrolled a total of 61 patients who underwent CT scanning for EAT volume and CAD and found no association between EAT volume and CAD based on the Gensini scores [28]. Despite the negative results, the majority of studies proved a significant positive correlation of EAT volume with CAD risk [20]. In the present study, we found that the correlation between EAT volume and CAD was non-linear in overall subjects. Further, by using a two-piecewise linear regression model, we calculated that the inflection point of the EAT-CAD correlation was 90 ml. Moreover, no studies assessed the relationship between EAT volume and the CAD risk factors in the South Asian population. Despite negative results of other investigated risk factors, we found linear correlations between EAT volume and BMI in men of the current Indian cohort. Our data showed EAT-CAD association was stronger when BMI was less than 30kg/m^2^ in men (Fig. 5), suggesting EAT volume as a better predictive maker for the disease in non-obese men. This phenomenon agrees with the fact that ectopic fat distribution or visceral fat deposition shows stronger correlations to CVDs [29,30]. Certainly, the unobstructed contiguity of EAT with the coronary arteries supports its stronger local effect than subcutaneous adipose tissue. In addition to the vicinity, the quantity and activity of EAT play more significant roles in CAD [20]. The complex mechanisms through which EAT causes CAD include adipocyte hypertrophy and hyperplasia, chronic inflammation, exaggerated innate immunity, endothelial damage, vascular remodeling, oxidative stress, lipotoxicity, and glucotoxicity [31].

### Comparison of data from different ethnic groups

Interestingly, the median EAT volume in the South Asian population (65.69ml) was similar to that of Japanese [32], but smaller than Chinese and European populations [24][33–36] and larger than populations with African ancestry [37,38] (Supp Table 5). Likewise, EAT volume had a larger effect size on CAD in Indian than in European [33–36]. In India, opposite to the global trend, the prevalence of central obesity is higher than general obesity and excessive visceral fat accumulation represents a stronger driver for obesity-associated CVD [7–9]. Indeed, in our analysis, EAT, a type of VAT, displayed a stronger association with CAD when BMI, the measure for general obesity, was less than 30kg/m^2^. The BMI cutoff of obesity for Asian and South Asian populations was recommended to be ≥ 25kg/m^2^ [39][40]. However, we only observed the influence of BMI on the association of EAT volume and CAD in men when a BMI cutoff of 30kg/m^2^ was used (Supp Table 6). For women, neither a cutoff of 25 nor 30kg/m^2^ indicated an impact of BMI on the EAT-CAD association (Supp Table 6). The data suggested that a sex-specific BMI definition of obesity might be considered for cardiovascular risk stratification. In fact, for a given BMI, women tend to have a higher body fat percentage than men.

### Sex-specific effects of EAT on CAD

We found a male-specific association of EAT volume with CAD in the current Indian cohort. Matched with our data, in the European population, men displayed larger EAT volume compared to women [41], and EAT volume was independently associated with coronary artery calcification in men but not in women [42]. In Japanese, EAT volume is one of the determinants for coronary artery bypass grafting (CABG) only in men [43], and EAT volume/body surface area was higher among male CAD patients, not in female patients [32]. The sex-specific difference in EAT volume or EAT-CAD association might be due to differences in lifestyle, hormonal level, and adipose tissue subtypes, cellular composition and molecular profile. In the current study cohort, for instance, male smokers were dramatically more than their female counterparts (n= 116 vs 2). Smoking induces multiple pathological changes in adipogenic differentiation, lipolysis and immune cell activation and infiltration in adipose tissue, contributing to CAD/atherosclerosis [44]. At the hormonal level, estrogen can promote insulin sensitivity, glucose metabolism and the fitness of adipocytes [45]. At the tissue level, the subcutaneous adipose tissue of females holds the better capability of fat deposition, preventing ectopic fat distribution around the organs including the heart [46]. Perivascular adipose tissue (PVAT) of women was shown to have features of brown or beige adipose tissue [47]. At the cellular and molecular levels, women showed a significantly smaller adipocyte size and lower lipoprotein lipase activity, which could reduce the free fatty acid production of EAT and therefore decrease the accumulation of lipids in the atherosclerotic plaques [43][48][49]. These reasons might partially explain a smaller EAT volume and insignificant EAT-CAD association in women of the current cohort.

### Strengths

Our study has several strengths. First, it was specifically conducted in the South Asian population, which, despite the large population number and high CVD incidence, represented an under-studied ethnic group. Our study was conducted in a sex-specific fashion allowing pinpoint the higher correlation between EAT volume and CAD risk in South Asian men than in women. The results might facilitate personalized medicine in this high-risk population. Second, for the first time, the association between EAT volume and CAD-related risk factors was investigated in a sex-specific manner among South Asians. Third, we not only use the generalized linear model to evaluate the relationship between EAT volume and CAD or its risk factors but also the generalized additive model to clarify the nonlinear relationship. A generalized additive model has obvious advantages in visualizing with non-linear correlation, smoothing the non-parametric values, and fitting a regression spline to the data. The use of a generalized additive model allowed us to better discover the real relationships between the exposure and the outcome. Fourth, although the cross-sectional design of the current study holds unavoidable potential confounding, we used strict statistical adjustment to minimize it. In addition, our subgroup analyses excluded the modification of EAT volume -CAD associations by traditional risk factors in the overall and woman subjects (Supp Fig. 1 and 2) but specified the effect of BMI on the association in men (Fig.5).

### Limitations

Certain limitations exist in our study. First, the paracardial adipose tissue, the other adipose depot in close contact with the heart and the coronary arteries, was not investigated in the current study. However, the epi- and para-cardial adipose volumes were highly correlated [50,51] and our result might also be suggestive of the effect of paracardial adipose volume on CAD as well as its risk factors. Second, body surface area (BSA) represents another good way to test the effect of EAT on CAD[52]. Unfortunately, we did not document the BSA. However, reports have shown similar results when EAT, EAT/BSA or EAT/BMI were used to test the effect of EAT on CAD [53]. Third, no guidelines have officially specified the attenuation threshold for EAT quantification. We specified the density range of –250 to –30 Hounsfield units to isolate adipose tissue, which was similar to many studies (Supp Table 5) but slightly differed from a few other studies [54][55]. However, as the EAT volume of our research subjects was measured with the same threshold range, our conclusion will remain the same. Fourth, this study is a cross-sectional study and thus provides only weak evidence between the exposure and the outcome. Further prospective follow-up studies are needed to verify the findings.

Finally, despite the advantage of population specificity in this study, the results may not be generalized to other ethnic groups.

## Conclusions

EAT volume was significantly and positively correlated with CAD and BMI in men, not in women, in the current South Asian population. The correlation of EAT volume with BMI was linear, but with CAD was non-linear in men. BMI affected the association of EAT volume with CAD risk when it is less than 30kg/m^2^, suggesting that EAT volume might be a better predictor or biomarker for CAD in non-obese men. These associations and sex differences should be considered in CAD prognostic models. Further well-designed, large-scale longitudinal studies are needed to confirm our conclusions and evaluate the underlying mechanisms of the association.

## Data Availability

All data produced in the present work are contained in the manuscript.

## Author contribution

ZC, DW, and TT conceptualized the study. ZC, RK, DW, TT, and CX designed the study and wrote the manuscript. CX, XG, LL, CL, AR and LFB ran analyses. RK, AY, and TB provided human data. MVS, AY, and TBS gave technical support and conceptual advice.

## Supplementary Materials

**Supplementary figure 1.**
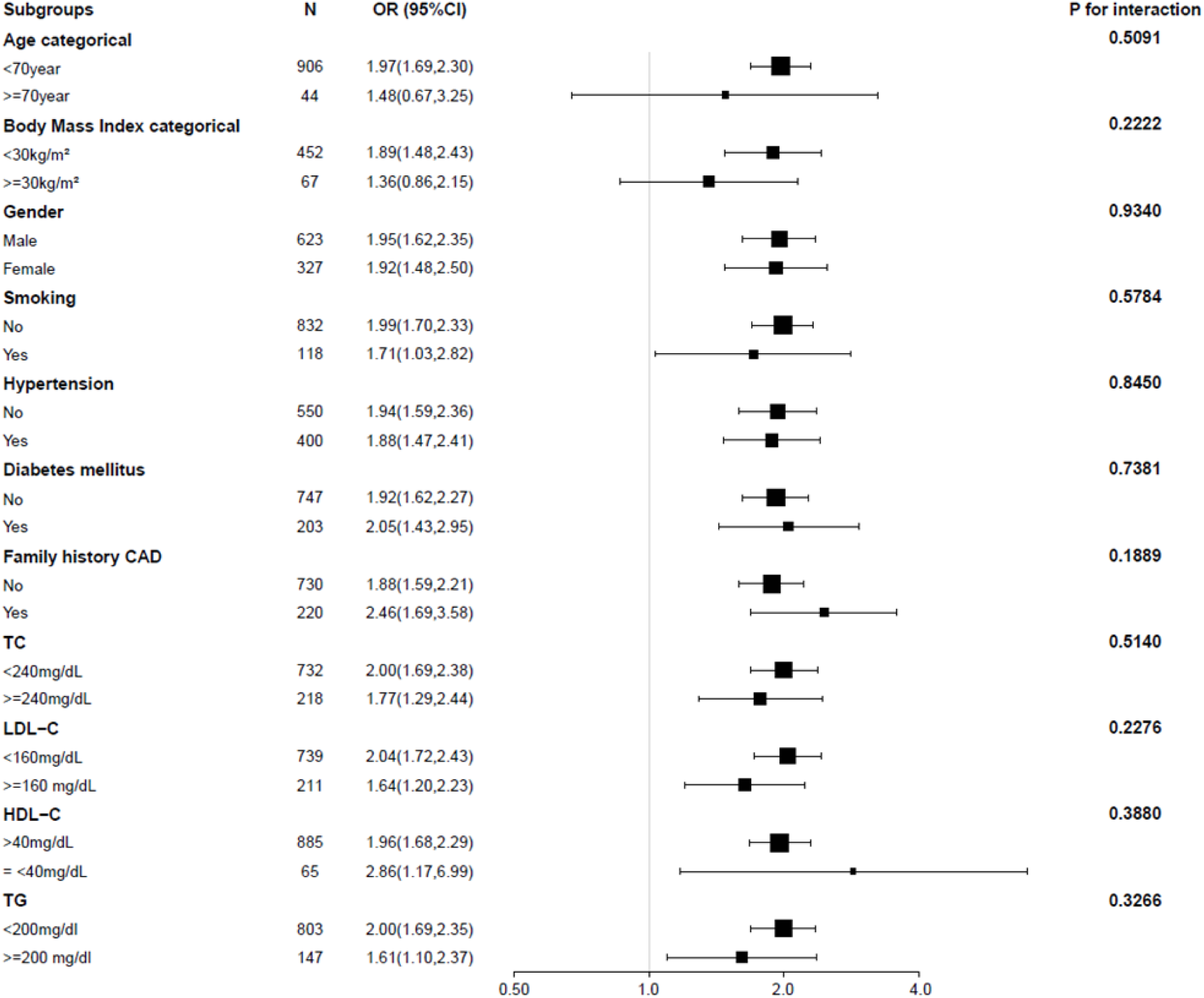
The association of EAT volume and CAD risk according to subgroups in the overall subjects. The effect of EAT volume on the risk of CAD was simillar in various subgroups. All analyses were adjusted for age, smoking, BMI, hypertension, diiabetes mellitus, famiily history of CAD, TG, TC, LDL-C and HDL-C. Riisk estiimates were for every 1-SD iincrease (31.0 ml} in EAT volume. Siize of squares are proportiional to CAD prevalence iin the specified subgroup. Horizontal error bars indicate 95% confidence iintervals. N=subjectsin each subgroup. OR, odds ratio.

**Supplementary figure 2.**
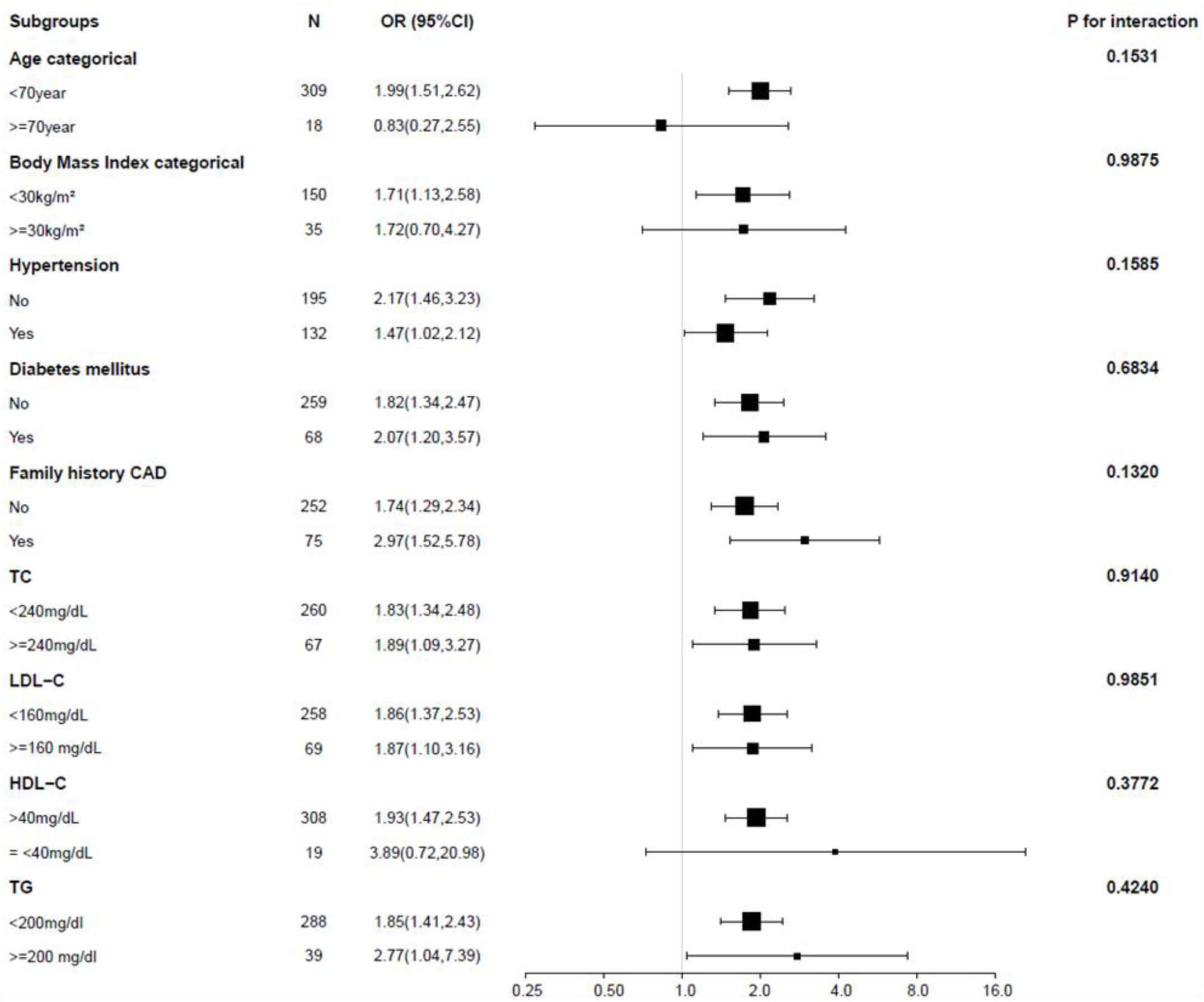
The association of EAT volume and CAD risk according to subgroups in women. The effect of EAT volume on the risk of CAD was similar in various subgroups. All analyses were adjusted for age, smokiing, BMI, hypertension, diiabetes mellitus, family history of CAD, TG, TC, LDL-C and HDL-C. Risk estiimates were for every 1-SD increase (28.9 ml) in EAT volume. Size of squares are proportional to CAD prevalence in the specified subgroup. Horizontal error bars indicate 95% confidence intervals. N=subjects in each subgroup. OR, odds ratio.

**Supplementary Table 1:** Quantile characteristics of overall subjects.

| Characteristic | Full cohort | Q1 (7.0-43.6) | Q2 (43.7-59.5) | Q3(59.7-82.5) | Q4(82.7-203.9) | P-value |
| --- | --- | --- | --- | --- | --- | --- |
| N | 950 | 238 | 237 | 237 | 238 |  |
| Age (years) | 51.1 ± 11.2 | 45.1 ± 12.4 | 51.1 ± 10.8 | 53.2 ± 9.7 | 55.0 ± 9.3 | <0.001 |
| Body Mass Index | 26.2 ± 3.5 | 25.1 ± 3.2 | 25.8 ± 2.8 | 26.7 ± 3.8 | 28.2 ± 3.9 | <0.001 |
| Gender |  |  |  |  |  | 0.04 |
| Male | 623 (65.6%) | 138 (58.0%) | 163 (68.8%) | 159 (67.1%) | 163 (68.5%) |  |
| Female | 327 (34.4%) | 100 (42.0%) | 74 (31.2%) | 78 (32.9%) | 75 (31.5%) |  |
| Smoking |  |  |  |  |  | 0.168 |
| No | 832 (87.6%) | 218 (91.6%) | 202 (85.2%) | 206 (86.9%) | 206 (86.6%) |  |
| Yes | 118 (12.4%) | 20 (8.4%) | 35 (14.8%) | 31 (13.1%) | 32 (13.4%) |  |
| Hypertension |  |  |  |  |  | <0.001 |
| No | 550 (57.9%) | 164 (68.9%) | 131 (55.3%) | 133 (56.1%) | 122 (51.3%) |  |
| Yes | 400 (42.1%) | 74 (31.1%) | 106 (44.7%) | 104 (43.9%) | 116 (48.7%) |  |
| Diabetes mellitus |  |  |  |  |  | 0.042 |
| No | 747 (78.6%) | 201 (84.5%) | 188 (79.3%) | 176 (74.3%) | 182 (76.5%) |  |
| Yes | 203 (21.4%) | 37 (15.5%) | 49 (20.7%) | 61 (25.7%) | 56 (23.5%) |  |
| Family history CAD |  |  |  |  |  | 0.299 |
| No | 730 (76.8%) | 192 (80.7%) | 175 (73.8%) | 178 (75.1%) | 185 (77.7%) |  |
| Yes | 220 (23.2%) | 46 (19.3%) | 62 (26.2%) | 59 (24.9%) | 53 (22.3%) |  |
| Total cholesterol |  |  |  |  |  | 0.476 |
| <200mg/dL | 732 (77.1%) | 186 (78.2%) | 189 (79.7%) | 181 (76.4%) | 176 (73.9%) |  |
| ≥200mg/dL | 218 (22.9%) | 52 (21.8%) | 48 (20.3%) | 56 (23.6%) | 62 (26.1%) |  |
| LDL-C |  |  |  |  |  | 0.191 |
| <100mg/dL | 739 (77.8%) | 191 (80.3%) | 192 (81.0%) | 180 (75.9%) | 176 (73.9%) |  |
| ≥100 mg/dL | 211 (22.2%) | 47 (19.7%) | 45 (19.0%) | 57 (24.1%) | 62 (26.1%) |  |
| HDL-C |  |  |  |  |  | 0.456 |
| >40mg/dL | 885 (93.2%) | 224 (94.1%) | 216 (91.1%) | 220 (92.8%) | 225 (94.5%) |  |
| ≤40mg/dL | 65 (6.8%) | 14 (5.9%) | 21 (8.9%) | 17 (7.2%) | 13 (5.5%) |  |
| Triglycerides |  |  |  |  |  | 0.196 |
| <150mg/dl | 803 (84.5%) | 211 (88.7%) | 197 (83.1%) | 200 (84.4%) | 195 (81.9%) |  |
| ≥150 mg/dl | 147 (15.5%) | 27 (11.3%) | 40 (16.9%) | 37 (15.6%) | 43 (18.1%) |  |
| CAD |  |  |  |  |  | <0.001 |
| No | 492 (51.8%) | 45 (76.3%) | 67 (59.3%) | 69 (52.7%) | 63 (40.6%) |  |
| Yes | 458 (48.2%) | 14 (23.7%) | 46 (40.7%) | 62 (47.3%) | 92 (59.4%) |  |

Values are N (Mean±SD) or N (%). EAT, epicardial adipose tissue; CAD, coronary artery disease; LDL-C, low density lipoprotein cholesterol; HDL-C, high density lipoprotein cholesterol; TG, triglycerides.

**Supplementary Table 2:** Quantile characteristics of men.

| Epicardial fat volume (mL) quartile |  | Q1 | Q2 | Q3 | Q4 | P-value |
| --- | --- | --- | --- | --- | --- | --- |
| N | 623 | 7.0-43.4 | 43.7-59.5 | 59.7-82.5 | 82.7-203.9 |  |
| EFV(mL) | 67.7 ± 31.8 | 31.9 ± 8.8 | 51.9 ± 4.3 | 70.8 ± 6.9 | 110.6 ± 23.7 | <0.001 |
| Age (years) | 50.6 ± 11.7 | 43.6 ± 13.7 | 50.3 ± 10.7 | 52.9 ± 9.9 | 54.7 ± 9.7 | <0.001 |
| Body Mass Index | 25.8 ± 3.1 | 24.7 ± 2.8 | 25.3 ± 2.5 | 26.3 ± 3.4 | 27.7 ± 3.1 | <0.001 |
| Gender |  |  |  |  |  | 0.429 |
| Male | 623 (100.0%) | 138 (100.0%) | 163 (100.0%) | 159 (100.0%) | 163 (100.0%) |  |
| Smoking |  |  |  |  |  | 0.534 |
| No | 507 (81.4%) | 118 (85.5%) | 129 (79.1%) | 128 (80.5%) | 132 (81.0%) |  |
| Yes | 116 (18.6%) | 20 (14.5%) | 34 (20.9%) | 31 (19.5%) | 31 (19.0%) |  |
| Hypertension |  |  |  |  |  | 0.079 |
| No | 355 (57.0%) | 92 (66.7%) | 89 (54.6%) | 86 (54.1%) | 88 (54.0%) |  |
| Yes | 268 (43.0%) | 46 (33.3%) | 74 (45.4%) | 73 (45.9%) | 75 (46.0%) |  |
| Diabetes mellitus |  |  |  |  |  | 0.187 |
| No | 488 (78.3%) | 117 (84.8%) | 127 (77.9%) | 119 (74.8%) | 125 (76.7%) |  |
| Yes | 135 (21.7%) | 21 (15.2%) | 36 (22.1%) | 40 (25.2%) | 38 (23.3%) |  |
| Family history CAD |  |  |  |  |  | 0.6 |
| No | 478 (76.7%) | 110 (79.7%) | 121 (74.2%) | 119 (74.8%) | 128 (78.5%) |  |
| Yes | 145 (23.3%) | 28 (20.3%) | 42 (25.8%) | 40 (25.2%) | 35 (21.5%) |  |
| Total cholesterol |  |  |  |  |  | 0.813 |
| <200mg/dL | 472 (75.8%) | 102 (73.9%) | 127 (77.9%) | 118 (74.2%) | 125 (76.7%) |  |
| ≥200mg/dL | 151 (24.2%) | 36 (26.1%) | 36 (22.1%) | 41 (25.8%) | 38 (23.3%) |  |
| LDL-C |  |  |  |  |  | 0.677 |
| <100mg/dL | 481 (77.2%) | 108 (78.3%) | 130 (79.8%) | 118 (74.2%) | 125 (76.7%) |  |
| ≥100 mg/dL | 142 (22.8%) | 30 (21.7%) | 33 (20.2%) | 41 (25.8%) | 38 (23.3%) |  |
| HDL-C |  |  |  |  |  | 0.164 |
| >40mg/dL | 577 (92.6%) | 131 (94.9%) | 145 (89.0%) | 147 (92.5%) | 154 (94.5%) |  |
| ≤40mg/dL | 46 (7.4%) | 7 (5.1%) | 18 (11.0%) | 12 (7.5%) | 9 (5.5%) |  |
| Triglycerides |  |  |  |  |  | 0.603 |
| <150mg/dl | 515 (82.7%) | 119 (86.2%) | 134 (82.2%) | 131 (82.4%) | 131 (80.4%) |  |
| ≥150 mg/dl | 108 (17.3%) | 19 (13.8%) | 29 (17.8%) | 28 (17.6%) | 32 (19.6%) |  |
| CAD |  |  |  |  |  | <0.001 |
| No | 166 (49.1%) | 33 (76.7%) | 47 (58.0%) | 49 (49.0%) | 37 (32.5%) |  |
| Yes | 172 (50.9%) | 10 (23.3%) | 34 (42.0%) | 51 (51.0%) | 77 (67.5%) |  |

Values are N (Mean±SD) or N (%). EAT, epicardial adipose tissue; CAD, coronary artery disease; LDL-C, low density lipoprotein cholesterol; HDL-C, high density lipoprotein cholesterol; TG, triglycerides.

**Supplementary Table 3:**
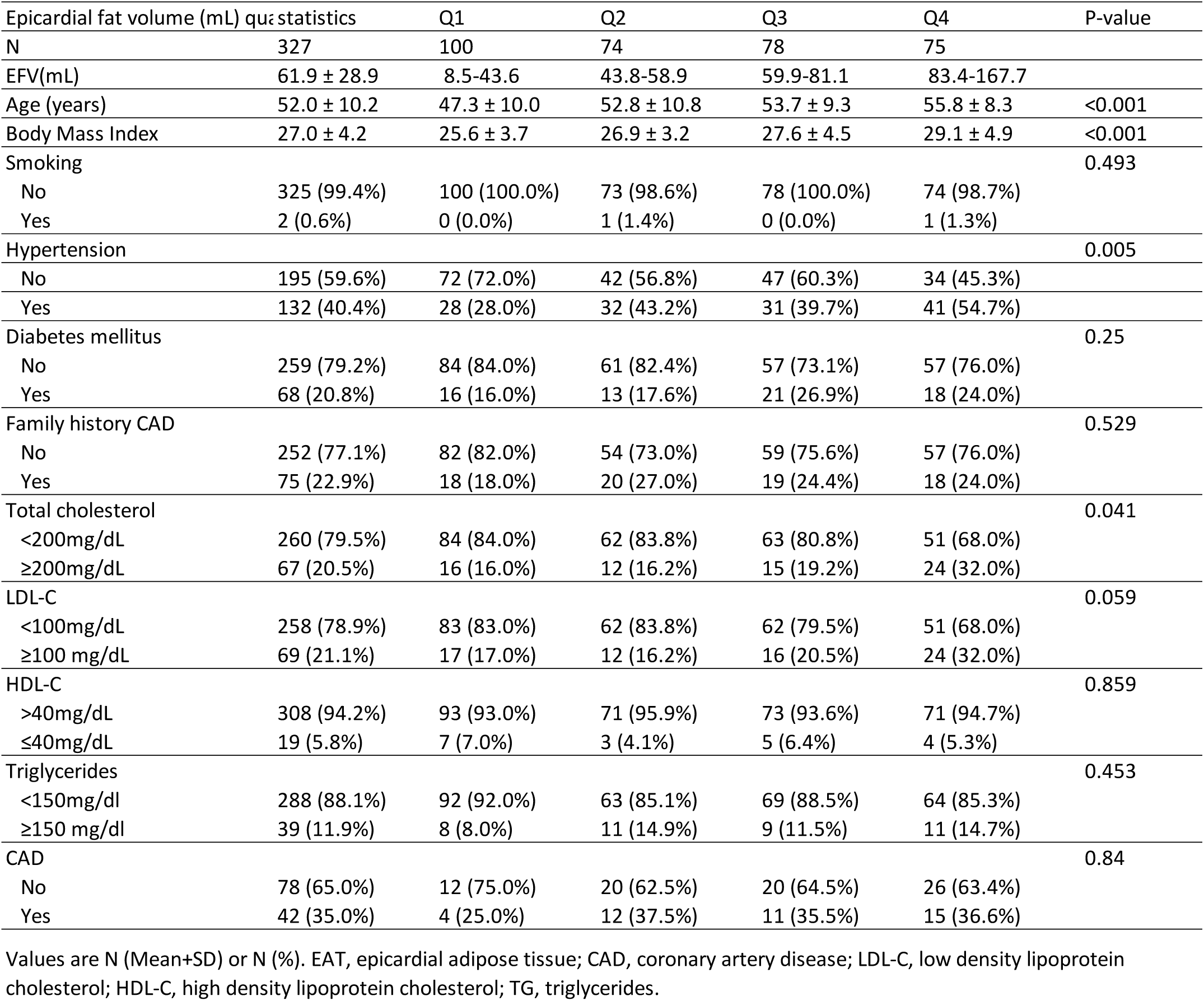
Quantile characteristics of women.

**Supplementary Table 4:**
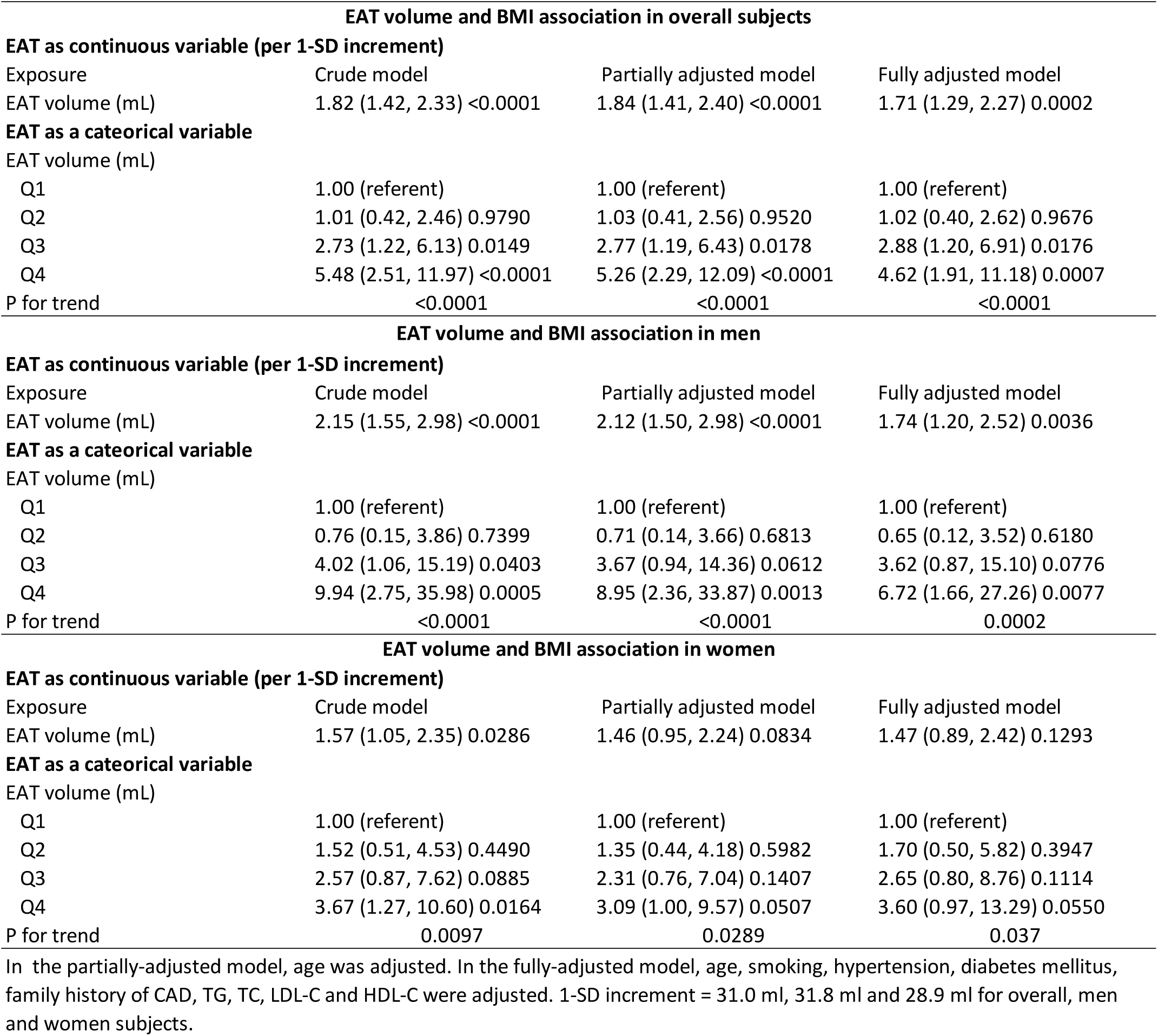
Multivariate analyses evaluating the association between EAT volume and BMI.

| **EAT volume and BMI association in overall subjects** |  |  |  |
| --- | --- | --- | --- |
| **EAT as continuous variable (per 1-SD increment)** |  |  |  |
| Exposure | Crude model | Partially adjusted model | Fully adjusted model |
| EAT volume (mL) | 1.82 (1.42, 2.33) <0.0001 | 1.84 (1.41, 2.40) <0.0001 | 1.71 (1.29, 2.27) 0.0002 |
| **EAT as a categorical variable** |  |  |  |
| EAT volume (mL) |  |  |  |
| Q1 | 1.00 (referent) | 1.00 (referent) | 1.00 (referent) |
| Q2 | 1.01 (0.42, 2.46) 0.9790 | 1.03 (0.41, 2.56) 0.9520 | 1.02 (0.40, 2.62) 0.9676 |
| Q3 | 2.73 (1.22, 6.13) 0.0149 | 2.77 (1.19, 6.43) 0.0178 | 2.88 (1.20, 6.91) 0.0176 |
| Q4 | 5.48 (2.51, 11.97) <0.0001 | 5.26 (2.29, 12.09) <0.0001 | 4.62 (1.91, 11.18) 0.0007 |
| P for trend | <0.0001 | <0.0001 | <0.0001 |
| **EAT volume and BMI association in men** |  |  |  |
| **EAT as continuous variable (per 1-SD increment)** |  |  |  |
| Exposure | Crude model | Partially adjusted model | Fully adjusted model |
| EAT volume (mL) | 2.15 (1.55, 2.98) <0.0001 | 2.12 (1.50, 2.98) <0.0001 | 1.74 (1.20, 2.52) 0.0036 |
| **EAT as a categorical variable** |  |  |  |
| EAT volume (mL) |  |  |  |
| Q1 | 1.00 (referent) | 1.00 (referent) | 1.00 (referent) |
| Q2 | 0.76 (0.15, 3.86) 0.7399 | 0.71 (0.14, 3.66) 0.6813 | 0.65 (0.12, 3.52) 0.6180 |
| Q3 | 4.02 (1.06, 15.19) 0.0403 | 3.67 (0.94, 14.36) 0.0612 | 3.62 (0.87, 15.10) 0.0776 |
| Q4 | 9.94 (2.75, 35.98) 0.0005 | 8.95 (2.36, 33.87) 0.0013 | 6.72 (1.66, 27.26) 0.0077 |
| P for trend | <0.0001 | <0.0001 | 0.0002 |
| **EAT volume and BMI association in women** |  |  |  |
| **EAT as continuous variable (per 1-SD increment)** |  |  |  |
| Exposure | Crude model | Partially adjusted model | Fully adjusted model |
| EAT volume (mL) | 1.57 (1.05, 2.35) 0.0286 | 1.46 (0.95, 2.24) 0.0834 | 1.47 (0.89, 2.42) 0.1293 |
| **EAT as a categorical variable** |  |  |  |
| EAT volume (mL) |  |  |  |
| Q1 | 1.00 (referent) | 1.00 (referent) | 1.00 (referent) |
| Q2 | 1.52 (0.51, 4.53) 0.4490 | 1.35 (0.44, 4.18) 0.5982 | 1.70 (0.50, 5.82) 0.3947 |
| Q3 | 2.57 (0.87, 7.62) 0.0885 | 2.31 (0.76, 7.04) 0.1407 | 2.65 (0.80, 8.76) 0.1114 |
| Q4 | 3.67 (1.27, 10.60) 0.0164 | 3.09 (1.00, 9.57) 0.0507 | 3.60 (0.97, 13.29) 0.0550 |
| P for trend | 0.0097 | 0.0289 | 0.037 |

In the partially-adjusted model, age was adjusted. In the fully-adjusted model, age, smoking, hypertension, diabetes mellitus, family history of CAD, TG, TC, LDL-C and HDL-C were adjusted. 1-SD increment = 31.0 ml, 31.8 ml and 28.9 ml for overall, men and women subjects.

**Supplementary Table 5:** EAT volume among different racial/ethnic groups.

| PMID | Ethnic population | Average EAT Volume | NCCT or CCTA | EAT threshold (HU) | Year of publication |
| --- | --- | --- | --- | --- | --- |
| 25109783 | African-Americans | 34.6 (23.1 to 47.3) cm<sup>3</sup> | NCCT | -190 to -30 | 2015 |
|  | Caucasians | 41.7 (32.4 to 57.4) cm<sup>3</sup> |  |  |  |
|  | Koreans | 43.5 (34.1 to 54.7) cm<sup>3</sup> |  |  |  |
|  | Japanese | 44.9 (33.2 to 56.7) cm<sup>4</sup> |  |  |  |
|  | Japanese-Americans | 51.9 (37.1 to 70.2) cm<sup>3</sup> |  |  |  |
| 24315112 | African American | 59 (39 to 84) cm<sup>3</sup> | CCTA | -195 to -45 | 2014 |
| 22963346 | Japanese | 65 ± 21 cm<sup>3</sup> (Women) | CCTA | -600 to -20 | 2012 |
|  |  | 80 ± 33 cm<sup>3</sup> (Men) |  |  |  |
| 32346001 | Australian | 72 ± 33 ml | CCTA | -190 to -30 | 2020 |
| 25896357 | European | 79.5 ± 34.3 cm<sup>3</sup> | NCCT | -195 to -45 | 2015 |
| 29233634 | American | 83.9 ± 38.0 cm<sup>3</sup> | NCCT | -190 to -30 | 2017 |
| 28954947 | Japanese | 86.4 ± 40.9 cm<sup>3</sup> | CCTA | -115 to -25 | 2018 |
| 33660521 | Chinese | 119.47 ± 36.56 cm<sup>3</sup> | NCCT | -190 to -30 | 2021 |
| Note: NCCT, non-contrast tomography angiography ; CCTA, contrast tomography angiography; HU, Hounsfield unit. |  |  |  |  |  |

**Supplementary Table 6:** EAT-CAD assocation in subgroups according to two BMI cutoffs.

| **Subgroups_All** | **N** | **OR (95%CI)** | **P for interaction** |
| --- | --- | --- | --- |
| Body Mass Index categorical |  |  | 0.2222 |
| <25kg/m<sup>2</sup> | 228 | 1.81(1.24,2.64) |  |
| >=25kg/m<sup>2</sup> | 291 | 1.66(1.28,2.16) |  |
| Body Mass Index categorical |  |  | 0.7158 |
| <30kg/m<sup>2</sup> | 452 | 1.89(1.48,2.43) |  |
| >=30kg/m<sup>2</sup> | 67 | 1.36(0.86,2.15) |  |
| **Subgroups_Men** | **N** | **OR (95%CI)** | **P for interaction** |
| Body Mass Index categorical |  |  | 0.8940 |
| <25kg/m<sup>2</sup> | 159 | 1.58(1.01,2.48) |  |
| >=25kg/m<sup>2</sup> | 175 | 1.64(1.17,2.31) |  |
| Body Mass Index categorical |  |  | **0.0381 \*** |
| <30kg/m<sup>2</sup> | 302 | 2.01(1.46,2.78) |  |
| >=30kg/m<sup>2</sup> | 32 | 0.99(0.56,1.75) |  |
| **Subgroups_Women** | **N** | **OR (95%CI)** | **P for interaction** |
| Body Mass Index categorical |  |  | 0.8940 |
| <25kg/m<sup>2</sup> | 69 | 2.32(1.16,4.64) |  |
| >=25kg/m<sup>2</sup> | 116 | 1.61(1.01,2.56) |  |
| Body Mass Index categorical |  |  | 0.9875 |
| <30kg/m<sup>2</sup> | 150 | 1.71(1.13,2.58) |  |
| >=30kg/m<sup>2</sup> | 35 | 1.72(0.70,4.27) |  |

